# Confounding factors of Alzheimer’s disease plasma biomarkers and their impact on clinical performance

**DOI:** 10.1101/2022.05.30.22275718

**Authors:** Alexa Pichet Binette, Shorena Janelidze, Nicholas Cullen, Jeffrey L. Dage, Randall J. Bateman, Henrik Zetterberg, Kaj Blennow, Erik Stomrud, Niklas Mattsson-Carlgren, Oskar Hansson

**Author notes:** Corresponding author: Oskar Hansson, Clinical Memory Research Unit, Memory Clinic, Skåne University Hospital, SE-20502 Malmö, Sweden.

## Abstract

**INTRODUCTION:** Plasma biomarkers will likely revolutionize the diagnostic work-up of Alzheimer’s disease (AD) globally. Before widespread use, we need to determine if confounding factors affect the levels of these biomarkers, and their clinical utility.

**METHODS:** Participants with plasma and CSF biomarkers, creatinine, body mass index (BMI), and medical history data were included (BioFINDER-1: n=748, BioFINDER-2: n=421). We measured beta-amyloid (Aβ42, Aβ40), phosphorylated tau (p-tau217, p-tau181), neurofilament light (NfL), and glial fibrillary acidic protein (GFAP).

**RESULTS:** In both cohorts, creatinine and BMI were the main factors associated with NfL, GFAP, and to a lesser extent with p-tau. However, adjustment for BMI and creatinine had only minor effects in models predicting either the corresponding levels in CSF or subsequent development of dementia.

**DISCUSSION:** Creatinine and BMI are related to certain plasma biomarkers levels, but they do not have clinically relevant confounding effects for the vast majority of individuals.

## 1. Introduction

Plasma biomarkers of pathologic events in Alzheimer’s disease (AD) and other neurodegenerative diseases are becoming more prevalent and increasingly accessible^1^. Plasma levels of beta-amyloid (Aβ) and phosphorylated tau (p-tau) reflect the two key pathological hallmarks of AD – brain amyloidosis and tau pathology, respectively – while plasma neurofilament light (NfL) is a marker of axonal degeneration that is elevated in many different neurodegenerative diseases^2-8^. Plasma glial fibrillary acidic protein (GFAP), a marker of glial activation, is also elevated early in the course of AD and has been increasingly studied^9,10^. Blood-based biomarkers can for instance discriminate controls from patients, and cognitively stable participants from those who convert to dementia, and they are related to the same markers measured in cerebrospinal fluid (CSF) or in the brain^11-15^. They are currently mainly available in research settings, but a lot of efforts are being devoted to making them more widely available, with potential use to screen participants for clinical trials and/or inform clinical practice in the near future^1,16^. However, determining if confounding factors affect the levels, and maybe even clinical utility, of these blood biomarkers is necessary before their widespread implementation. In particular, recent evidence suggests that reduced kidney function might be associated with increased plasma biomarkers concentrations^17,18^, and higher body mass index (BMI) with lower blood NfL levels^18,19^. In this study, we set out to study the effects of potential confounding factors on several state-of-the-art blood-based biomarkers for AD and neurodegeneration, including p-tau217, p-tau181 and GFAP, as well as Aβ42/Aβ40 and NfL, in two independent cohorts. We first investigated associations between blood-based biomarkers levels and common comorbidities and medication use, as well as factors that are proxies to blood volume (body mass index [BMI]) and kidney function (plasma creatinine). This allowed us to identify key potential confounding factors of plasma biomarkers concentrations. More importantly, we studied whether the performance of the blood-based biomarkers was improved when adjusting for potential confounding factors to determine whether such adjustments are needed in future clinical settings. For this, we studied whether the potential confounding factors either influenced i) the associations between individual plasma biomarkers and their CSF counterparts, or (ii) the ability of the plasma biomarkers to predict conversion to AD dementia or all-cause dementia in non-demented individuals.

## 2. Methods

### 2.1. Participants

Participants from the two independent prospective Swedish BioFINDER-1 (NCT01208675) and BioFINDER-2 (NCT03174938) cohort studies were included. All participants were recruited at Skåne University Hospital and the Hospital of Ängelholm, Sweden and cover the full spectrum of AD, ranging from older adults with intact cognition or subjective cognitive decline, mild cognitive impairment (MCI), and dementia. Both studies were approved by the Regional Ethics Committee in Lund, Sweden. All participants gave written informed consent to participate. BioFINDER-1 participants were enrolled between January 2010 and December 2014 and were followed longitudinally for up to 8 years. Baseline data and conversion status to AD dementia or any dementia was used from BioFINDER-1. BioFINDER-2 is an ongoing longitudinal study that was launched in 2017, which does not overlap with BioFINDER-1. For the current study, baseline data was included from participants recruited between April 2017 and April 2021. All cognitively unimpaired participants, from both BioFINDER-1 and 2, were recruited from a community-based study, the Malmö Diet and Cancer study.

The main inclusion criteria were to be 60 years or older in BioFINDER-1 or 40 years and older in BioFINDER-2, being fluent in Swedish, having mini-mental state evaluation (MMSE) between 27 and 30 for cognitively normal participants and between 24 and 30 for MCI. Exclusion criteria were having significant unstable systemic illness, neurological or psychiatric illness, significant alcohol or substance misuse, or refusing lumbar puncture or neuroimaging. MCI diagnostic differed slightly between studies. In BioFINDER-1, MCI status was determined after an extensive neuropsychological assessment if participant performed worse than one to two standard deviations on various tests^20^. In BioFINDER-2, MCI diagnosis was established if participants performed below 1.5 standard deviation from norms on at least one domain from an extensive neuropsychological battery examining verbal, episodic memory, visuospatial ability, and attention/executive domains^14^. Dementia diagnosis was determined by consensus of memory clinic physicians and neuropsychologist. For AD dementia, diagnosis was based on the criteria from the Diagnostic and Statistical Manual of Mental Disorders Fifth Edition and if positive on Aβ biomarkers based on the updated NIA-AA criteria for AD ^21^. BioFINDER-2 further included a cohort of non-AD dementias and neurodegenerative disorders in which fulfilment of criteria for dementia or major neurocognitive disorder could be due to frontotemporal dementia, Parkinson’s disease, vascular dementia, dementia with Lewy bodies, progressive supranuclear palsy, multiple system atrophy, corticobasal syndrome or primary progressive aphasia. Greater detail of both studies has been described previously^7,14,22^.

### 2.2. Medical history

In both cohorts, information on medical history and medications were recorded at the baseline visit. The information was retrieved from both the medical records of each participant and questionnaires answered by the participant and/or his/her informant.

### 2.3. Creatinine and body mass index measurements

All participants had data available for plasma creatinine and BMI. Creatinine levels were measured in plasma taken from a blood draw at the baseline visit. Blood was sampled in the morning (not after fasting) and each sample was analyzed at Skåne University Hospital. The CREP2 Cobas 501 (2016-12, V13.0) or CREP2 Cobas 701 (2018-03, V10.0) analytical unit from Roche Diagnostics were used to measure creatinine (reported in μmol/l). In BioFINDER-1, the average time between the blood draw at baseline to measure creatinine and the blood draw/lumbar puncture to measure biomarkers was 12 days ± 36 days, with 6 participants (<1%) exceeding a 6-month difference. In BioFINDER-2, it was 51 days ± 53 days, with no one exceeding 6-month difference.

To calculate BMI, height and weight were taken at baseline visit for every participant.

### 2.4. Plasma and cerebrospinal fluid biomarkers

Biomarkers analyzed in plasma and CSF included Aβ42, Aβ40, p-tau217, p-tau181, NfL, and GFAP. All biomarker concentrations were measured from blood and CSF samples taken the same day in the morning, not after fasting.

#### 2.4.1. Aβ42 and Aβ40

In BioFINDER-1, plasma Aβ42 and Aβ40 were measured using Elecsys immunoassays on a Cobas e 601 module (Roche Diagnostics) as described previously^3^. In BioFINDER-2, plasma Aβ42 and Aβ40 were measured using the liquid chromatography mass-spectrometry based technology from Araclon Biotech, as described recently^23^. In both cohorts, CSF Aβ42 and Aβ40 were measured using the Elecsys immunoassays (Roche Diagnostics)^24^.

For a subset of BioFINDER-1 participants (n=635), plasma Aβ42 and Aβ40 was also measured using the immunoprecipitation-coupled mass spectrometry developed at Washington University (WashU-IP-MS), as described previously^5^. In head-to-head comparisons with 8 other plasma Aβ assays, the IP-MS-WashU assay was recently shown as the best one to identify participants classified as being Aβ-positive on CSF and on PET^25^. We validated the main plasma Aβ findings with the WashU-IP-MS assays, and all results can be found in Supplementary Material.

#### 2.4.2. p-tau217 and p-tau181

In both BioFINDER-1 and BioFINDER-2, plasma as well as CSF p-tau217 was measured using immunoassays on the Meso-Scale Discovery platform developed by Lilly Research Laboratories as described previously^20,26^. The calibration of the assays differed between cohorts, which explains the different range of values. In BioFINDER-1 the assays were calibrated with a synthetic p-tau217 peptide, while in BioFINDER-2 a recombinant tau protein phosphorylated *in vitro* was used. All details have been described recently^23^.

For p-tau181, plasma assays differed between cohorts. In BioFINDER-1 the assay from Lilly Research Laboratories based on the Meso-Scale Discovery platform was used, as described recently^7^, and was available for 570 participants out of 748. In BioFINDER-2, the Simoa assay developed at the University of Gothenburg was used^2^. CSF p-tau181 was measured using the Elecsys immunoassays (Roche Diagnostics)^24,27^ in both cohorts.

In both cohorts, some participants had plasma p-tau217 and p-tau181 levels below the detection level of the assay (56 participants [7.5%] in BioFINDER-1 and 52 participants [12%] in BioFINDER-2 for p-tau217; 44 participants [7.7%] in BioFINDER-1 and 10 participants [2.4%] in BioFINDER-2 for p-tau181). In those cases, values were imputed by the lowest detection level of the assay. We also repeated the analyses excluding the participants with very low values and all results remained the same.

#### 2.4.3. NfL

In both cohorts, plasma NfL concentrations were measured using the commercially available Single molecule array (Simoa)^28,29^, and CSF NfL was measured using the Elecsys immunoassay (Roche Diagnostics).

#### 2.4.4. GFAP

In BioFINDER-2, plasma GFAP was measured with Simoa GFAP Discovery kits for SR-X, as previously described^30^. For plasma GFAP in BioFINDER-1 and CSF GFAP in both cohorts, the Elecsys immunoassay (Roche Diagnostics) was used^10^.

### 2.5. Statistical analysis

The two cohorts were analyzed separately. First, we used linear regression models (including age and sex as covariates) to investigate the associations of each plasma marker (dependent variable) with creatinine, BMI, comorbidities, and medication use separately, to identify the most important potential confounders for further analyses. For associations that were significant, we further adjusted the model for global Aβ-PET SUVR (flutemetamol)^14^ to evaluate if the associations between plasma biomarkers and potential confounding factors were independent of existing pathology (e.g. prodromal AD has been shown to be directly associated with lower BMI, which is likely to be a consequence of the disease^31^). Second, we wanted to evaluate if the identified factors influenced the associations between each biomarker measured in plasma and its CSF counterpart. For that, we compared the plasma coefficient between two regression models. The first model only included basic co-variates (CSF level ∼ plasma level + age + sex). The second model further included the potential confounders as covariates (e.g. CSF level ∼ plasma level + age + sex + creatinine + BMI). To determine whether the plasma coefficients were significantly different between the two models, we used a non-parametric procedure: we bootstrapped each model over 10 000 iterations and subtracted the plasma coefficients from all iterations between the two models. We then used the 95% confidence interval (CI) of this difference to assess if plasma estimates differed between models. Third, we wanted to evaluate whether the key confounders influenced the plasma estimate in assessing conversion to either AD dementia or all-cause dementia over a four-year period^20^, using logistic regression. This was only done in BioFINDER-1, where longitudinal data was available. Non-dementia converters with less than four years of follow-up were censored. For analyses related to conversion to AD dementia, individuals who converted to another type of dementia were excluded. We used a similar approach as for the plasma-CSF comparisons, with a bootstrap procedure to compare odds ratio of plasma biomarkers from logistic models that discriminated between participants who remained stable vs. those who converted to dementia. The base logistic regression model included plasma biomarker, age and sex and the second model further included key confounders as covariates. Standardized coefficients are reported for all analyses, so that different models can be compared. For data visualization, the raw biomarkers values are shown, to display accurately the range of values of the different assays used. To further investigate the effect of adjusting for potential confounding factors at the individual level, we also calculated the relative change (in %) in predicting the dependent variable from the base model vs. the model including confounding factors for each participant (more details and results in Figure S3).

All analyses were performed on R version 4.0.5 using the packages ABA (“Automated Biomarker Analyses) version 1.0.0^15^ for linear regressions, Boot version 1.3-27 for bootstrapping and pROC version 1.17.0.1 for logistic regression.

## 3. Results

### 3.1. Participants

Two-thirds of BioFINDER-1 participants were cognitively normal and the remaining third were MCI participants (Table 1). Diagnoses were more varied in BioFINDER-2, where 56%, 23%, and 21% were diagnosed as cognitively normal, MCI or AD dementia or non-AD neurodegenerative diseases (52 with AD, 3 with Parkinson’s disease, 6 with progressive supranuclear palsy, 6 with Lewy body dementia, 1 with corticobasal syndrome, 7 with behavioral variant frontotemporal dementia, 3 with multiple system atrophy, 3 with primary progressive aphasia, 7 with vascular dementia, 1 with unspecified cause of dementia) respectively. Both cohorts were balanced between men and women and creatinine levels and BMI were similar between cohorts. Generally, both cohorts presented similar frequency of comorbidities and medication use, with the exception that more participants in BioFINDER-1 had hypertension, hyperlipidemia, and a previous stroke (Table 1). Since all plasma assays apart from NfL differed between cohorts, each cohort was analyzed separately (see Table S1 for average plasma and CSF biomarkers levels). The main biomarkers of interest were the Aβ42/Aβ40 ratio, p-tau217, NfL and GFAP. Results related to Aβ42 and Aβ40 used individually, and p-tau181 can be found in Supplementary material.

**Table 1.**
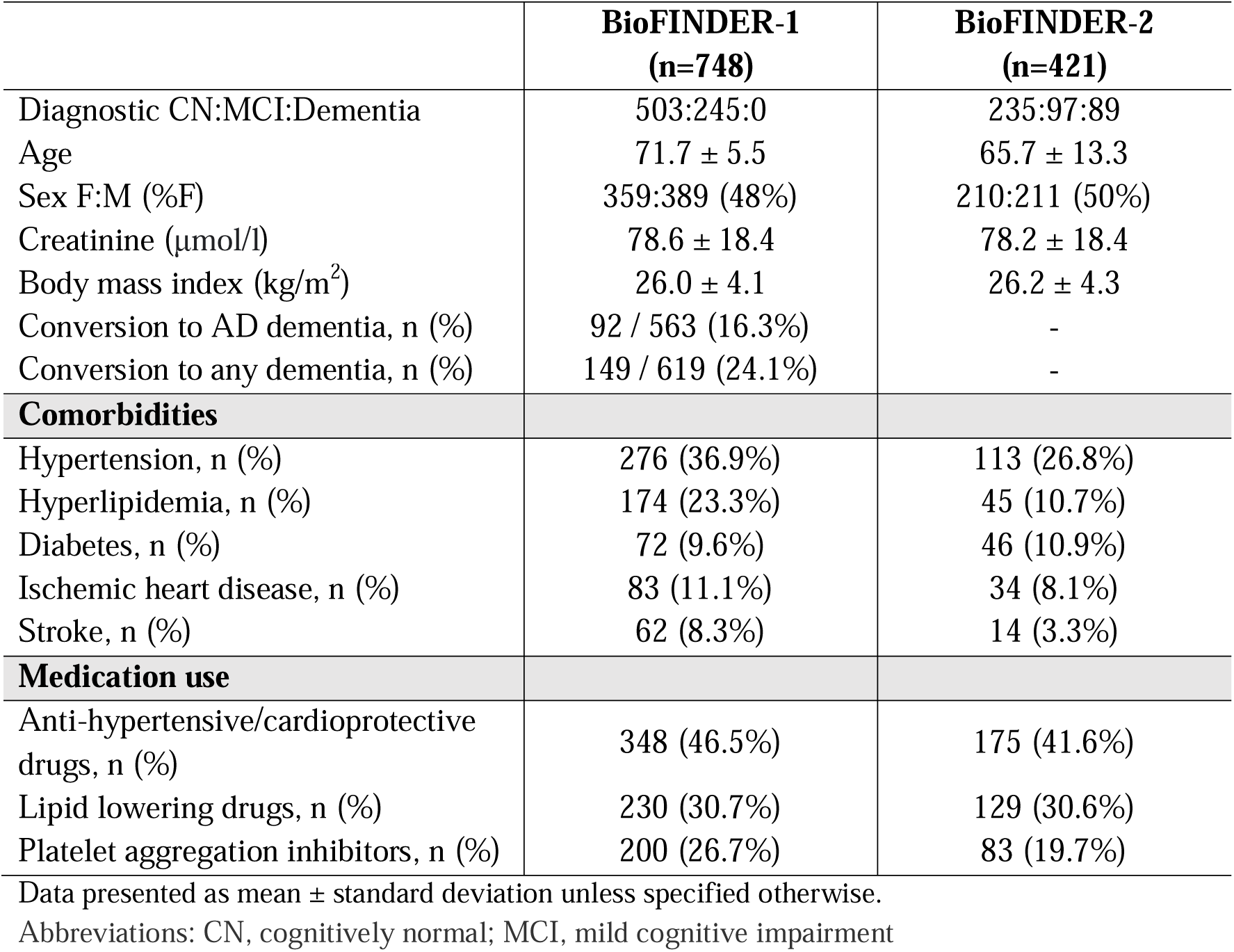
Demographics of BioFINDER-1 and BioFINDER-2 cohorts.

### 3.2. Associations of creatinine, body mass index and comorbidities with plasma biomarkers

Of all evaluated variables, creatinine and BMI were the main factors associated with different plasma biomarkers concentrations (Figure 1). In both cohorts, creatinine level was positively associated with NfL and GFAP levels. In BioFINDER-1 only, higher creatinine was also associated with higher p-tau217 and p-tau181 (Figure S1). In both cohorts, BMI was negatively associated with NfL, GFAP, p-tau217 and p-tau181 levels. Further, creatinine and BMI were associated with Aβ42 and Aβ40 alone (Figure S1), but not with the Aβ42/Aβ40 ratio. Across both cohorts, there were no consistent effect of different comorbidities and medication use on plasma levels of Aβ42/40, p-tau217, p-tau181, NfL or GFAP (Figure 1 and S1). Rather, comorbidities or medication use were more related to levels of Aβ42 and Aβ40 alone, but not their ratio. Hypertension, diabetes, taking anti-hypertensive/ cardioprotective medication or lipid lowering medication were related to both higher plasma Aβ42 and Aβ40 levels (Figure S1), as reported previously^32^. In BioFINDER-1, a subset of participants had plasma Aβ levels measured with the WashU-IP-MS assay (n=635), in which the same effects on Aβ42 and Aβ40 alone, but none on the Aβ42/Aβ40 ratio, were corroborated (Figure S2).

**Figure 1.**
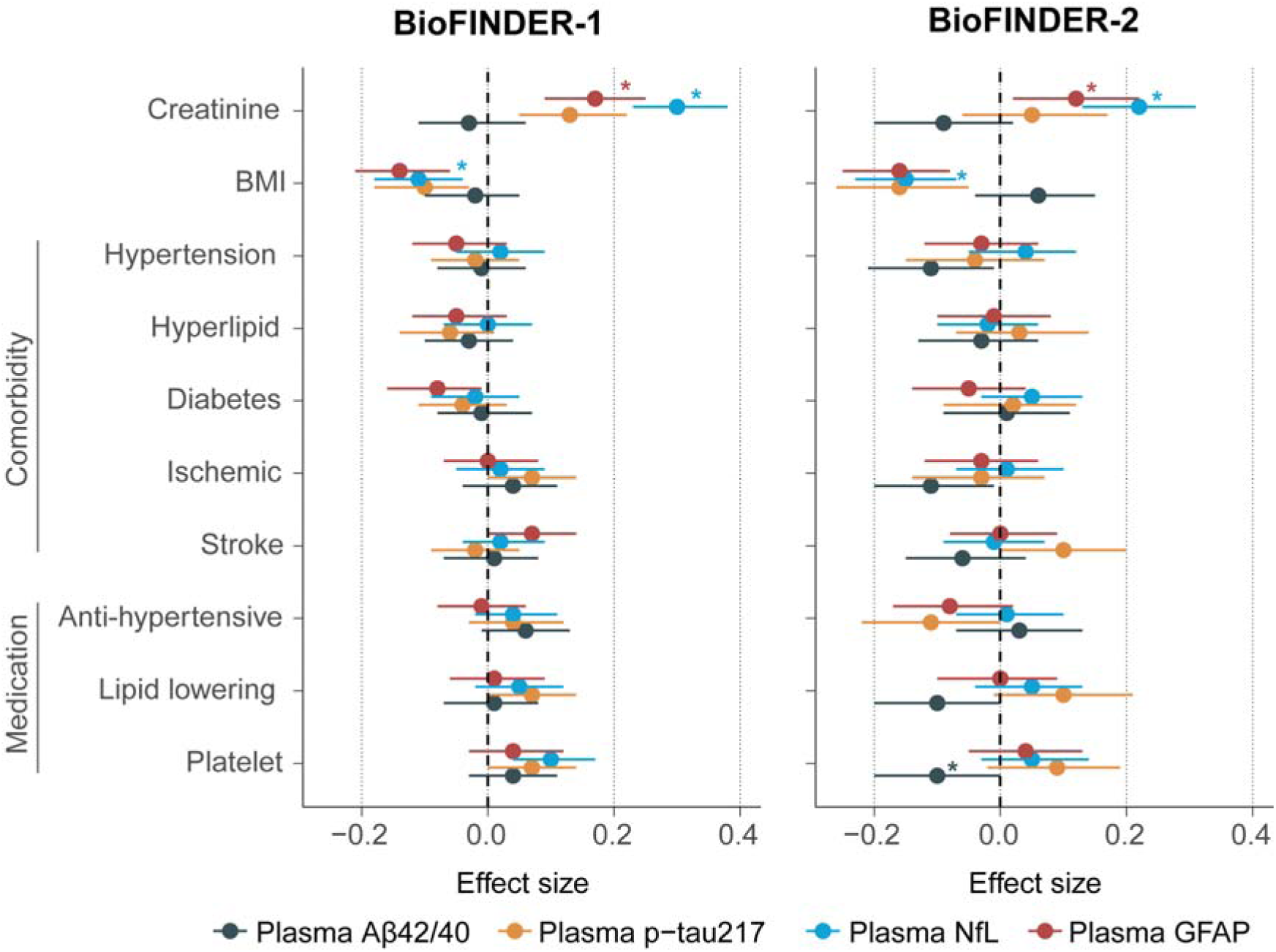
Associations between creatinine, body mass index, comorbidities and medication use with plasma biomarkers. Standardized beta coefficients and 95% confidence interval from linear regression models adjusted for age and sex with plasma biomarkers in BioFINDER-1 (left) and BioFINDER-2 (right). The Aβ42/40 ratio was reversed so that the effect size moves in the same direction for each biomarker. As such, higher value on each biomarker is more abnormal (towards AD). Stars indicate that the association remained significant when further adjusting for flutemetamol global Aβ-PET SUVR. In BioFINDER-2, all associations also remained significant if adding temporal meta-ROI tau-PET SUVR from [^18^F]RO948 as a covariate. Abbreviations: Aβ, beta-amyloid; BMI, body mass index; CSF, cerebrospinal fluid, GFAP, glial fibrillary acidic protein; NfL, neurofilament light; p-tau217, phosphorylated tau 217

We further examined if the significant associations between potential confounders and plasma biomarkers were independent of brain pathology by adding global Aβ-PET SUVR as a covariate. The associations of creatinine with NfL and GFAP were unchanged. However, BMI was no longer associated with plasma biomarkers, except for plasma NfL (Figure 1). All associations with Aβ42 and Aβ40 alone remained unchanged (Figure S1). These results suggest that the association of plasma biomarker concentrations with creatinine are likely due to a confounding effect on plasma concentrations (i.e., lower kidney function resulting in somewhat higher biomarker concentrations especially for NfL and GFAP). On the other hand, associations with BMI might be due partly to blood volume diluting plasma biomarkers concentrations in the case of NfL but to general disease progression for other markers (i.e., a lower BMI in individuals with prodromal AD induced by the disease^31,33^). Overall, given that the strongest and consistent associations with plasma biomarkers were with creatinine and BMI, subsequent analyses focused on these two confounders.

### 3.3. Effect of creatinine and BMI on plasma-to-CSF associations of AD biomarkers

Next, we investigated whether accounting for creatinine and BMI influenced the associations between plasma and CSF concentrations of each biomarker. Generally, p-tau217, followed by NfL showed the greatest correlations between the concentrations in plasma and the concentrations in CSF of the same biomarker (Figure 2 and Table 2).

**Table 2.** Plasma biomarker coefficients related to corresponding cerebrospinal fluid biomarker levels.

| | Aβ42/40 | | p-tau217 | | NfL | | GFAP | |
| --- | --- | --- | --- | --- | --- | --- | --- | --- |
| | $\beta$ | R<sup>2</sup> | $\beta$ | R<sup>2</sup> | $\beta$ | R<sup>2</sup> | $\beta$ | R<sup>2</sup> |
| **BioFINDER-1** |  |  |  |  |  |  |  |  |
| Base model | 0.41 | 0.18 | 0.66 | 0.43 | **0.49** | 0.27 | **0.31** | 0.13 |
| Model including creatinine and BMI | 0.41 | 0.19 | 0.66 | 0.45 | **0.54** | 0.29 | **0.33** | 0.13 |
| **BioFINDER-2** |  |  |  |  |  |  |  |  |
| Base model | **0.34** | 0.26 | 0.51 | 0.33 | **0.64** | 0.49 | 0.40 | 0.43 |
| Model including creatinine and BMI | **0.35** | 0.31 | 0.49 | 0.36 | **0.68** | 0.50 | 0.41 | 0.43 |

Significance of all plasma coefficients and of all models is $p < 0.001$ .
Abbreviations: Aβ , beta-amyloid; BMI, body mass index; GFAP, glial fibrillary acidic protein; NfL, neurofilament light; p-tau217, phosphorylated tau 217

**Figure 2.**
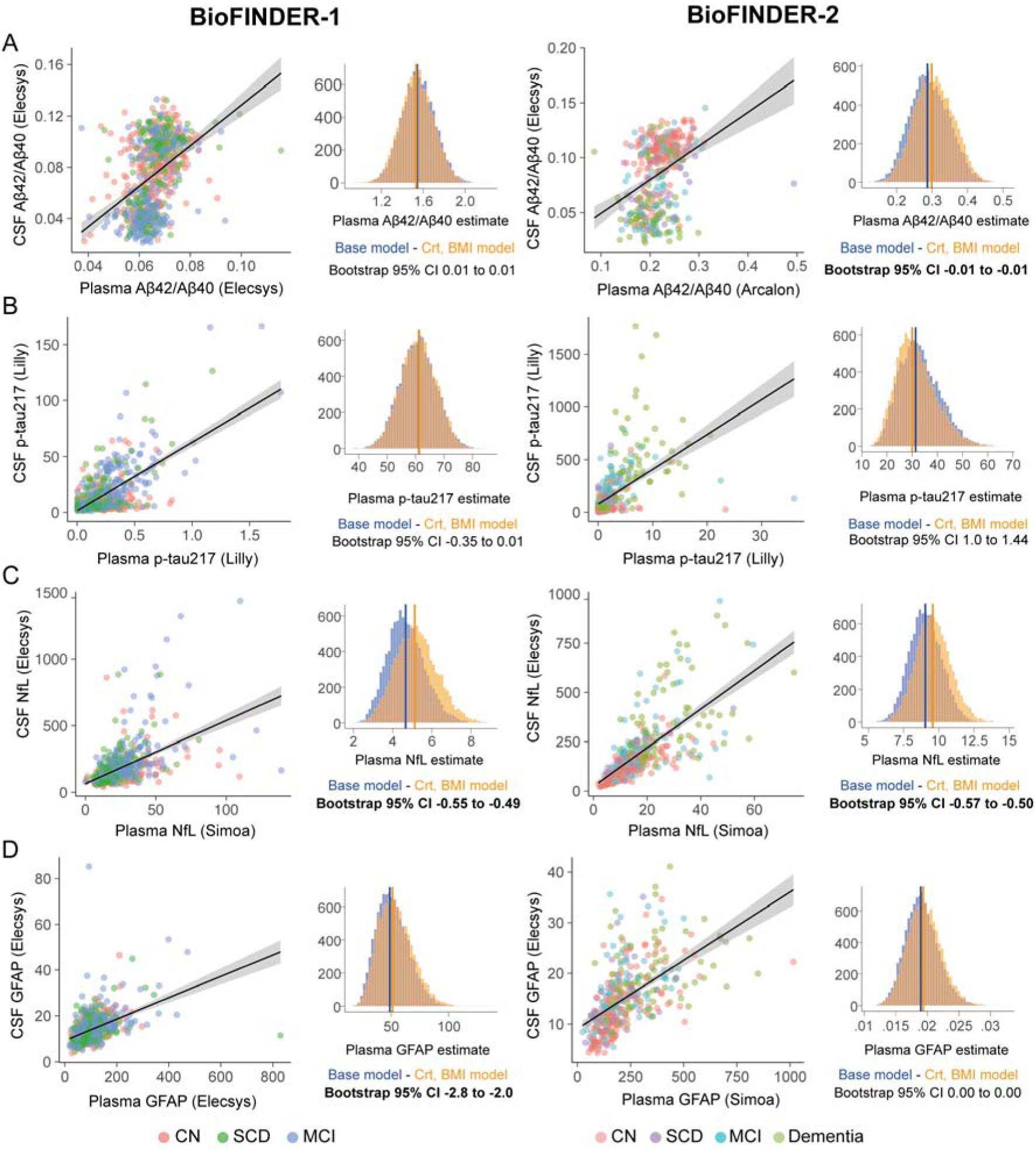
Associations between plasma and corresponding cerebrospinal fluid biomarkers. Scatter plots depict bivariate associations between plasma and CSF levels for Aβ42/40 ratio (A), p-tau217 (B), NfL (C) and GFAP (D) in BioFINDER-1 on the left-hand side and BioFINDER-2 on the right-hand side. To compare plasma coefficients between the base model and the one including creatinine and BMI as covariates, we generated 10 000 bootstrap samples of both models, shown in the histograms. Significant difference between models was based on the 95% confidence interval difference of the difference of plasma estimates between models. Abbreviations: Aβ, beta-amyloid; BMI, body mass index; Crt: creatinine, CSF, cerebrospinal fluid, GFAP, glial fibrillary acidic protein; NfL, neurofilament light; p-tau217, phosphorylated tau 217

In both cohorts, NfL was the only biomarker for which accounting for creatinine and BMI consistently improved the plasma coefficient (by 6 to 10%) in relation to CSF when using plasma concentration as independent variable (Figure 2C, Table 2). When inverting the associations (using CSF concentration as the independent variable), no improvements were seen in CSF estimates when accounting for creatinine and BMI, showing that these two confounders only affect plasma, and not CSF, concentrations. At the individual level, we evaluated the relative change in percentage in predicting CSF NfL in the basic model vs. the models further including creatinine and BMI as covariates. Generally, the plasma NfL model adjusted for creatinine improved the predictions of CSF NfL concentrations in participants with high creatinine values, but worsened the predictions in subjects with low creatinine values (R=0.44 (BioFINDER-1) and R=0.28 (BioFINDER-2) from associations between relative change between models and creatinine; Figure S3), while there was no or only minor effect of BMI (Figure S3). However, in general, the relative change between models across participants was modest, with an average of 0% and standard deviation of 13%. The number of participants where the adjusted plasma NfL model improved the prediction of the CSF NfL concentrations was just slightly higher (50% in BioFINDER-1 and 51% in BF-2) compared to the number of cases where the model worsened the prediction (49% in BioFINDER-1 and 44% in BioFINDER-2) when compared to the basic plasma NfL model.

Plasma GFAP estimates were improved when the CSF measure was accounted for creatinine and BMI in BioFINDER-1 by 6%, but no effect was seen in BioFINDER-2 (Figure 2D, Table 2). Regarding the associations in Aβ biomarkers, the strongest effects of adjusting for creatinine and BMI were seen on Aβ42 and Aβ40 alone (Table S2 and Figure S4, Table S3 for WashU-IP-MS assays), while effects on the Aβ42/40 ratio were more limited, with improvements between 2 and 3% (Figure 2A, Table 2).

### 3.4. Effect of creatinine, and body mass index on plasma biomarkers to estimate subsequent conversion to dementia in non-demented participants

In BioFINDER-1, long-term longitudinal data was available, and participants had follow-up visits. In this cohort we assessed whether accounting for creatinine and BMI influenced the plasma estimate to discriminate participants who remained stable vs. those who converted to dementia within a 4-year period. ROC curves from models assessing conversion to AD dementia using plasma Aβ42/40, p-tau217 and GFAP, and conversion to all-cause dementia using NfL are shown in Figure 3, and results from all models are shown in Table 3. When further including creatinine and BMI in models, plasma p-tau217 odds ratio in discriminating AD dementia converters from stable participants, and plasma NfL odds ratio in discriminating all-cause dementia converters from stable participants were improved by 4.5% (Table 3). Similar results as with p-tau217 was also observed with p-tau181 (Table S4). Although plasma estimates were significantly improved when accounting for creatinine and BMI, the discriminative accuracies between models were virtually the same, with a maximum change in AUC of 0.01. Looking at the individual level, the relative changes between basic model vs. when including creatinine and BMI for p-tau217 and NfL was lower than the changes seen in the plasma-CSF associations, with a standard deviation of 5%. Again, slightly more participants had better outcome prediction when including creatinine and BMI in logistic regressions (44% and 48% for p-tau217 and NfL respectively) compared to the number of cases where the model adjusted for confounding factors was worse than the basic model (33% and 40% for p-tau217 and NfL respectively, Figure S5). Here improvements in the models adjusted for creatinine and BMI were seen in participants with higher creatinine levels and higher BMI for both biomarkers (R=0.33 and 0.36 for associations between relative change between models and creatinine, and R=0.29 for same associations with BMI, Figure S5).

**Table 3.**
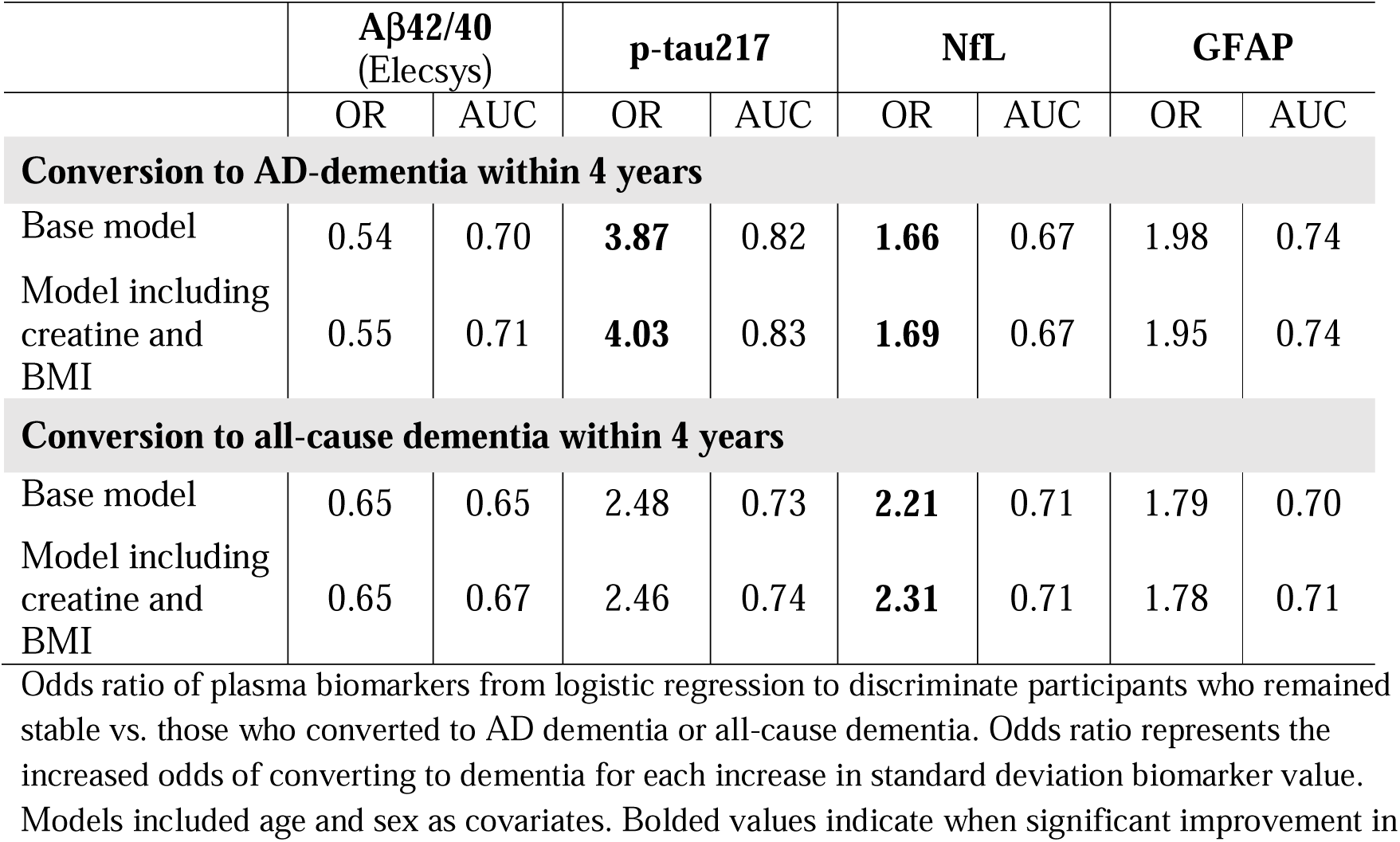
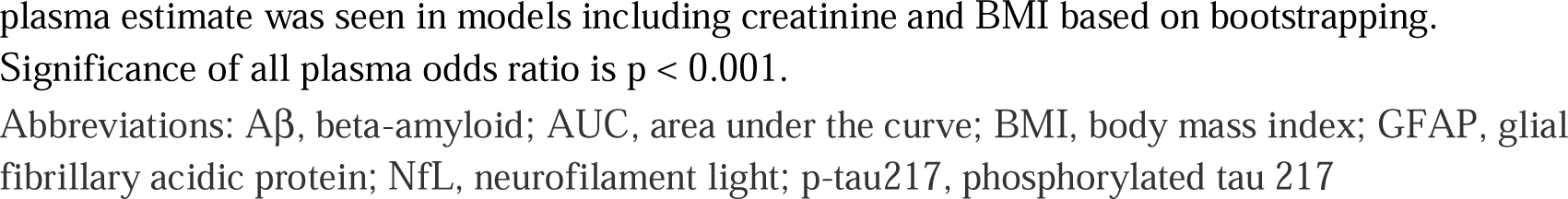
Plasma estimates related to conversion to dementia from logistic regressions in BioFINDER-1.

| | Aβ42/40<br>(Elecsys) | | p-tau217 | | NfL | | GFAP | |
| --- | --- | --- | --- | --- | --- | --- | --- | --- |
|  | OR | AUC | OR | AUC | OR | AUC | OR | AUC |
| **Conversion to AD-dementia within 4 years** |  |  |  |  |  |  |  |  |
| Base model | 0.54 | 0.70 | **3.87** | 0.82 | **1.66** | 0.67 | 1.98 | 0.74 |
| Model including creatine and BMI | 0.55 | 0.71 | **4.03** | 0.83 | **1.69** | 0.67 | 1.95 | 0.74 |
| **Conversion to all-cause dementia within 4 years** |  |  |  |  |  |  |  |  |
| Base model | 0.65 | 0.65 | 2.48 | 0.73 | **2.21** | 0.71 | 1.79 | 0.70 |
| Model including creatine and BMI | 0.65 | 0.67 | 2.46 | 0.74 | **2.31** | 0.71 | 1.78 | 0.71 |

Odds ratio of plasma biomarkers from logistic regression to discriminate participants who remained stable vs. those who converted to AD dementia or all-cause dementia. Odds ratio represents the increased odds of converting to dementia for each increase in standard deviation biomarker value. Models included age and sex as covariates. Bolded values indicate when significant improvement in
plasma estimate was seen in models including creatinine and BMI based on bootstrapping.
Significance of all plasma odds ratio is $p < 0.001$ .
Abbreviations: Aβ , beta-amyloid; AUC, area under the curve; BMI, body mass index; GFAP, glial fibrillary acidic protein; NfL, neurofilament light; p-tau217, phosphorylated tau 217

**Figure 3.**
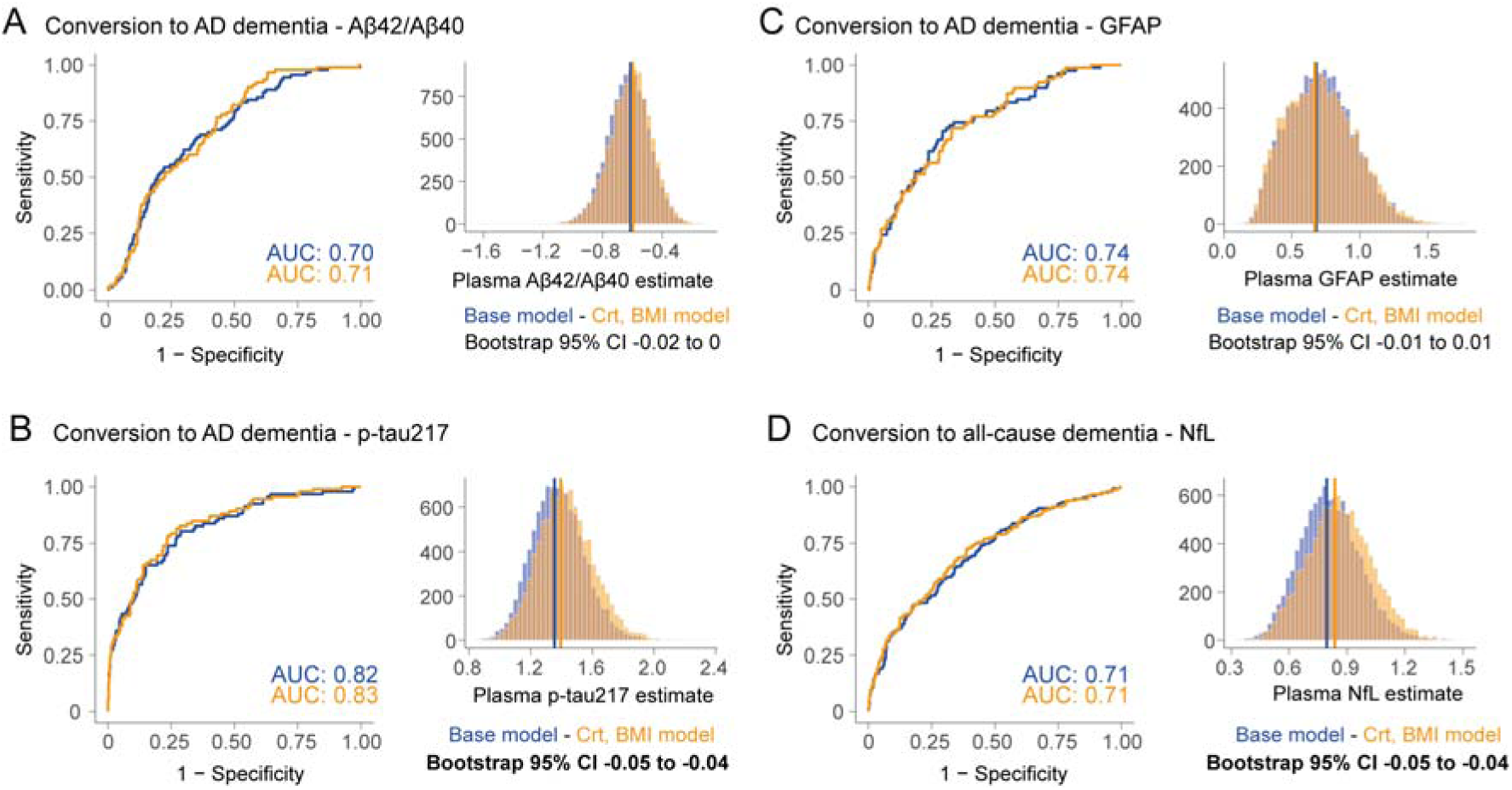
Plasma biomarkers on assessing conversion to AD or all-cause dementia in non-demented participants. ROC curves showing accuracy to discriminate between BioFINDER-1 participants who remained cognitively normal vs those who progressed to AD dementia based on plasma Aβ42/Aβ40 ratio (A), p-tau217 (B) and GFAP (C), and to discriminate participants who remained cognitively normal vs those who progressed to all-cause dementia based on plasma NfL (D). Results from logistic regression including plasma levels, age and sex are sown in the blue curve and models including creatinine and BMI as additional covariates in the orange curve. Plasma estimates between both models were then compared using bootstrapping as shown in the histograms and as done previously.

## 4. Discussion

We investigated whether comorbidities, medication use, plasma creatinine and BMI – measures routinely assessed in clinical settings – influenced key plasma AD markers in two large and independent cohorts. We studied whether such measures affected plasma markers levels, and whether they influenced the associations between plasma and CSF markers and the ability of plasma markers to predict future dementia. Our focus was on Aβ42/Aβ40 ratio, p-tau217, NfL and GFAP, and supplementary analyses included Aβ42, Aβ40 and p-tau181. Creatinine and BMI were related to concentrations of plasma NfL, GFAP, and to a lesser extent p-tau217 and 181, but not the Aβ42/Aβ40 ratio. However, including creatinine and BMI in models aimed towards clinical outcomes provided only very modest effect in improving plasma estimates when predicting either i) the corresponding concentrations in CSF or ii) subsequent development of dementia in non-demented individuals.

We investigated a comprehensive array of comorbidities and medication use, but creatinine and BMI were the two measures being consistently associated with plasma concentrations of several AD-related biomarkers. Higher creatinine levels and lower BMI were related to greater plasma concentrations of certain markers (especially plasma NfL and GFAP, and to a lesser extent p-tau). The directionality of such results might be explained by lower blood filtration (and thus higher creatinine) being associated with greater levels of biomarkers, and that greater blood volume, due to higher BMI, dilutes biomarkers levels in plasma. Further, most associations between creatinine and plasma biomarkers seem to be independent of pathology in the brain as measured by PET, which was not the case for most associations with BMI. Apart from NfL, the relations between lower BMI and more abnormal plasma biomarkers concentrations are likely due to loss of weight associated with prodromal AD^31,34^. Consequently, plasma creatinine is likely to have confounding effect on plasma biomarkers related to blood filtration, but the effect of BMI might not solely be a result of a confounding effect of higher blood volume in subjects with higher BMI, except for NfL which was associated with BMI even when adjusting for Aβ-PET. Our results are in line with previous studies that showed associations between creatinine, kidney function or BMI and plasma NfL and Aβ^18,19,35^. We also found associations with these confounding factors and plasma Aβ42 or Aβ40 alone, but not with the ratio Aβ42/Aβ40 which was consistent in two cohorts studied here using three different Aβ assays. Generally, all associations detected with either Aβ42 or Aβ40 where not present with the Aβ42/40 ratio, further highlighting the ratio as a measure of choice less affected by confounding factors.

An important novelty of the study resides in extending analysis to evaluate to which extent the main confounding factors (i.e., kidney function and BMI) affected the clinical performance of the blood-based biomarkers, i.e., their ability to predict i) their corresponding concentrations in CSF (which should be more closely related to the levels in the brain) and ii) clinical progression to dementia (which is a very clinically relevant outcome). First, we reported correlations between the concentrations in plasma and CSF for each marker, which were especially strong for NfL and p-tau217^16,36^. Given that the highest effect of creatinine and BMI were seen on plasma NfL concentrations, it is perhaps not surprising that creatinine and BMI improved the plasma NfL estimates related to NfL concentrations in the CSF, which was consistent in both cohorts. We note, however, that the improvement in the magnitudes of plasma estimates and R^2^ values were very modest, when assessed at the group level (Figure 2C and Table 2). Further, when assessed at the individual level very few participants exhibited clearly improved prediction of the CSF NfL concentration when plasma NfL was adjusted for either plasma creatinine or BMI (Figure S3). Several studies have shown that plasma biomarkers can be used to predict subsequent development of AD dementia as well as all-cause dementia^7,15,20^, however, no study has yet determined whether adjustment for kidney function or BMI might improve such predictions. We found that estimates for p-tau217 (when predicting AD dementia) and NfL (when predicting all-cause dementia) were improved when creatinine and BMI were considered. However, those improvements were even smaller than those seen in the plasma-CSF associations. The accuracies to differentiate stable participants from the ones who converted to dementia were virtually unchanged by including creatinine and BMI, with a change of only 0.01 in AUCs between models (Figure 3 and Table 3). Similarly, at the individual level very few participants exhibited improved prediction of dementia when adjusting for creatinine or BMI (Figure S5). We therefore propose that BMI and creatinine play a minor role in predictive value of plasma biomarkers for the vast majority of individuals assessed. Still, the classification accuracy of plasma biomarkers and basic demographics alone to predict clinical progression is in line with other studies, with p-tau217 being the best marker for AD dementia^20,37^.

The study has various strengths and limitations. We relied on two large, independent, and well-characterized cohorts. While the assays were not always the same between the two cohorts, most of the results were nevertheless consistent across the two cohorts. As the plasma Aβ assays relied on different technology between cohorts (immunoassays and mass spectrometry), we validated the main results using the currently best mass spectrometry assays available in a large subset of participants^25^. Results are reflective of two cohorts in which the majority of participants have BMI or creatinine levels within normal range. For instance, across both cohorts, around 15% of participants have BMI above 30, and 6% of women had creatinine levels above 90 μmol/l and 11% of men had creatinine levels above 105 μmol/l, taken as general cut-points for high values. Participants did not present with severe kidney diseases, which in itself had been related to greater risk of cognitive impairment^38^ and shown to affect plasma biomarkers levels^18^. Given that we only studied baseline levels of plasma biomarkers, it will be important to evaluate how fluctuations in plasma creatinine levels might influence longitudinal changes in plasma biomarker concentrations, because when used as outcomes in trials rather small changes in biomarker concentrations over time (e.g., 10-15%) might be considered relevant and thereby more susceptible to minor variations. Also, our current focus was on using measures that are routinely assessed clinically, like plasma creatinine. Future studies may also include other proxies of kidney function, such as cystatin-C, which may allow a more precise estimate of the glomerular filtration rate^39,40^.

Taken together, we showed that plasma creatinine levels and BMI were related to axonal degeneration marker NfL in plasma, and to a lesser extent to GFAP and p-tau proteins. The current study suggests that creatinine and BMI only have minor effects on improving the performance of AD-relevant plasma biomarkers. Consequently, diagnostic and prognostic algorithms for AD based on plasma biomarkers generally do not need to be adjusted for creatinine levels or BMI, even though these two factors have been the two most relevant confounding variables for plasma AD biomarkers identified until now.

## Data Availability

Anonymized data can be shared to qualified academic researchers after request for the purpose of replicating procedures and results presented in the study. Data transfer must be in agreement with EU legislation regarding general data protection regulation and decisions by the Ethical Review Board of Sweden and Region Skane, which should be regulated in a data transfer agreement.

## List of abbreviations

Aβ: beta-amyloid
AD: Alzheimer’s disease
AUC: area under the curve
BMI: body mass index
CI: confidence interval
CSF: cerebrospinal fluid
GFAP: glial fibrillary acidic protein
NfL: neurofilament light
p-tau: phosphorylated tau

## 5. Acknowledgements

We would like to acknowledge all of the BioFINDER team members as well as participants in the study and their family members for their dedication and patience.

