## Supplementary material for "Confounding factors of Alzheimer’s disease plasma biomarkers and their impact on clinical performance"

Table S1. Plasma and CSF biomarkers levels

Figure S1. Associations between creatinine, BMI, comorbidities, and medication use with plasma and CSF Aβ40, Aβ42 and p-tau181

Figure S2. Associations between creatinine, BMI, comorbidities, and medication use with WashU-IP-MS plasma Aβ markers

Figure S3. Effect of adjusting NfL plasma-CSF associations for creatinine and BMI at the individual level

Table S2. Plasma Aβ40, Aβ42 and p-tau181biomarker coefficients related to corresponding cerebrospinal fluid biomarker levels

Figure S4. Associations between plasma Aβ40, Aβ42 and p-tau181 and corresponding cerebrospinal fluid corresponding biomarkers

Table S3. WashU-IP-MS plasma Aβ markers coefficients related to corresponding cerebrospinal fluid biomarker levels

Table S4. Plasma Aβ42 and p-tau181 estimates related to conversion to dementia from logistic regressions in BioFINDER-1

Figure S5. Effect of adjusting p-tau217 and NfL models for creatinine and BMI in relation to conversion to dementia at the individual level

**Table S1. Plasma and CSF biomarkers levels**

| | **BioFINDER-1 (n=748)** | **BioFINDER-2 (n=421)** |
|---|---|---|
| **Plasma markers** | | |
| Aβ42 (pg/ml)[1] | 31.8 ± 5.0 | 64.6 ± 14.7 |
| Aβ40 (pg/ml)[1] | 484.0 ± 69.3 | 299.0 ± 59.1 |
| Aβ42/40[1] | 0.07 ± 0.01 | 0.22 ± 0.04 |
| p-tau217 (pg/ml)[1] | 0.22 ± 0.20 | 2.5 ± 3.8 |
| p-tau181 (pg/ml) | 3.4 ± 1.6[2] | 8.1 ± 8.2 |
| NfL (pg/ml) | 24.1 ± 14.5 | 16.9 ± 10.7 |
| GFAP (pg/ml)[1] | 104.8 ± 64.6 | 226.9 ± 141.7 |
| **Cerebrospinal fluid markers** | | |
| Aβ42 (pg/ml) | 1294.0 ± 684.3 | 1607.4 ± 813.1 |
| Aβ40 (ng/ml) | 17.31 ± 5.3 | 18.9 ± 5.9 |
| Aβ42/40 | 0.08 ± 0.03 | 0.09 ± 0.03 |
| p-tau217 (pg/ml)[1] | 14.4 ± 18.1 | 159.8 ± 232.1 |
| p-tau181 (pg/ml) | 22.3 ± 11.0 | 21.8 ± 13.0 |
| NfL (pg/ml) | 178.9 ± 137.9 | 189.4 ± 152.5 |
| GFAP (ng/ml) | 14.3 ± 9.7 | 15.2 ± 6.6 |

Data presented as mean ± standard deviation unless specified otherwise. Apart from NfL, plasma assays differed between cohorts. For CSF biomarkers, all assays were the same between cohorts, except for p-tau217.

[1] Assays differed between BioFINDER-1 and BioFINDER-2, see Methods for details.

[2] Only available for 570 participants

Aβ = beta-amyloid; p-tau = phosphorylated tau; NfL = neurofilament light; GFAP=glial fibrillary acidic protein

**Figure S1. Associations between creatinine, BMI, comorbidities and medication use with plasma Aβ40, Aβ42 and p-tau181**

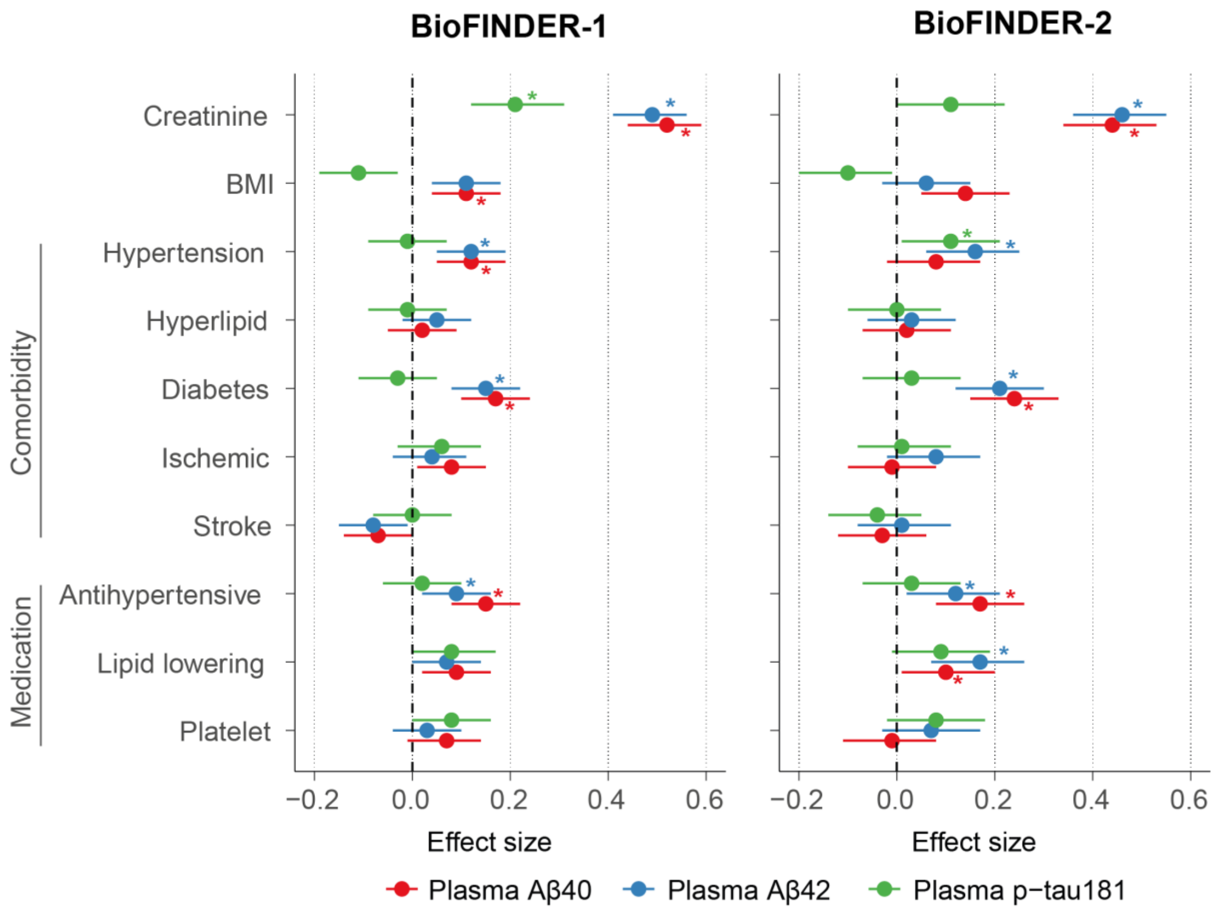


Standardized beta coefficients and 95% confidence interval from linear regression models adjusted for age and sex. The star denotes that the association remained significant when further adjusting for flutemetamol global Aβ-PET SUVR. In BioFINDER-2, all associations also remained significant if adding temporal meta-ROI tau-PET SUVR from [$^{18}$F]RO948 as a covariate.

Abbreviations: Aβ, beta-amyloid; BMI, body mass index; CSF, cerebrospinal fluid, p-tau181, phosphorylated tau 181

**Figure S2. Associations between creatinine, BMI, comorbidities, and medication use with WashU-IP-MS plasma Aβ markers**

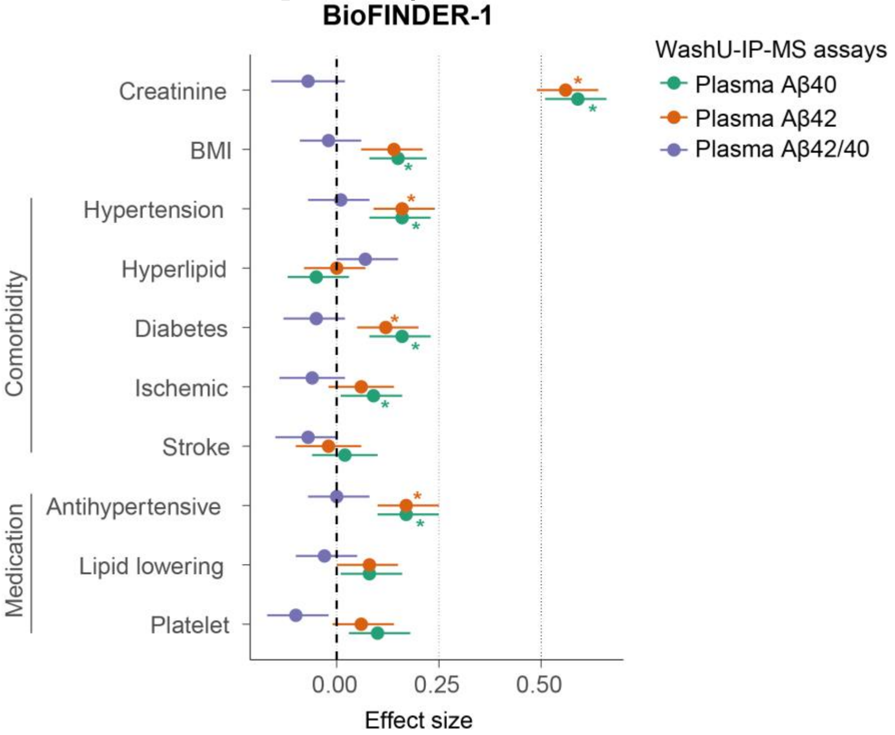


Standardized beta coefficients and 95% confidence interval from linear regression models adjusted for age and sex. The star denotes that the association remained significant when further adjusting for flutemetamol global Aβ-PET SUVR.

Abbreviations: Aβ, beta-amyloid; BMI, body mass index

**Figure S3. Effect of adjusting NfL plasma-CSF associations for creatinine and BMI at the individual level**

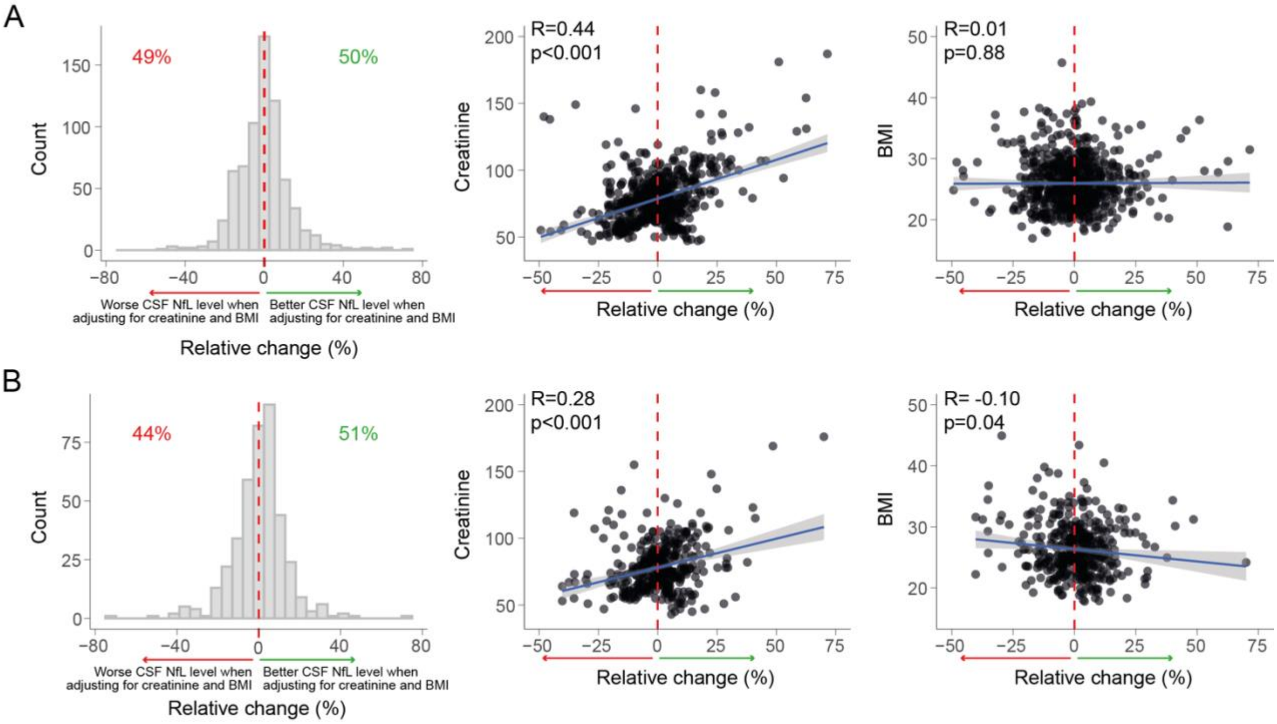


Relative change in NfL plasma-CSF association when accounting for creatinine and BMI at the individual level in BioFINDER-1 (A) and BioFINDER-2 (B). Relative change at the individual level was calculated as 100*( | predicted CSF value from basic model - observed CSF | - | predicted CSF value from model with creatinine and BMI - observed CSF | ) /observed CSF. As such, positive change (right to the 0% dotted red line) favors the model including creatinine and BMI (better prediction of CSF NfL concentration). Conversely, a negative change (left to the 0% dotted red line) favors the basic model (better prediction of CSF NfL concentration without including creatinine and BMI). Relations between relative change at the individual level and creatinine levels or BMI are shown in scatter plots. Abbreviations: BMI, body mass index; NfL, neurofilament light

**Table S2. Plasma Aβ40, Aβ42 and p-tau181biomarker coefficients related to corresponding cerebrospinal fluid biomarker levels**

| | **Aβ40** | | **Aβ42** | | **p-tau181** | |
|---|---|---|---|---|---|---|
| | β | $R^2$ | β | $R^2$ | β | $R^2$ |
| **BioFINDER-1** | | | | | | |
| Bivariate model | **0.12** | 0.03 | **0.30** | 0.10 | 0.48 | 0.26 |
| Model including creatinine and BMI | **0.15** | 0.03 | **0.34** | 0.10 | 0.49 | 0.29 |
| **BioFINDER-2** | | | | | | |
| Bivariate model | -0.06 | 0.03 | **0.23** | 0.08 | 0.19 | 0.15 |
| Model including creatinine and BMI | -0.05 | 0.03 | **0.25** | 0.09 | 0.18 | 0.19 |

Standardized beta coefficients of plasma biomarkers from models assessing CSF (dependent variable) and plasma biomarkers levels adjusted for age and sex, or from models further including creatinine and BMI as covariates. Adjusted $R^2$ of models are also reported. Bolded values indicate when significant improvement in plasma estimate was seen in models including creatinine and BMI based on bootstrapping. Significance of all plasma coefficients and of all models is $p < 0.001$, except for Aβ40 in BioFinder-2 where association between plasma and CSF was not significant.

Abbreviations: Aβ, beta-amyloid; BMI, body mass index; p-tau181 phosphorylated tau 181

**Figure S4. Associations between plasma Aβ40, Aβ42 and p-tau181 and corresponding cerebrospinal fluid corresponding biomarkers**

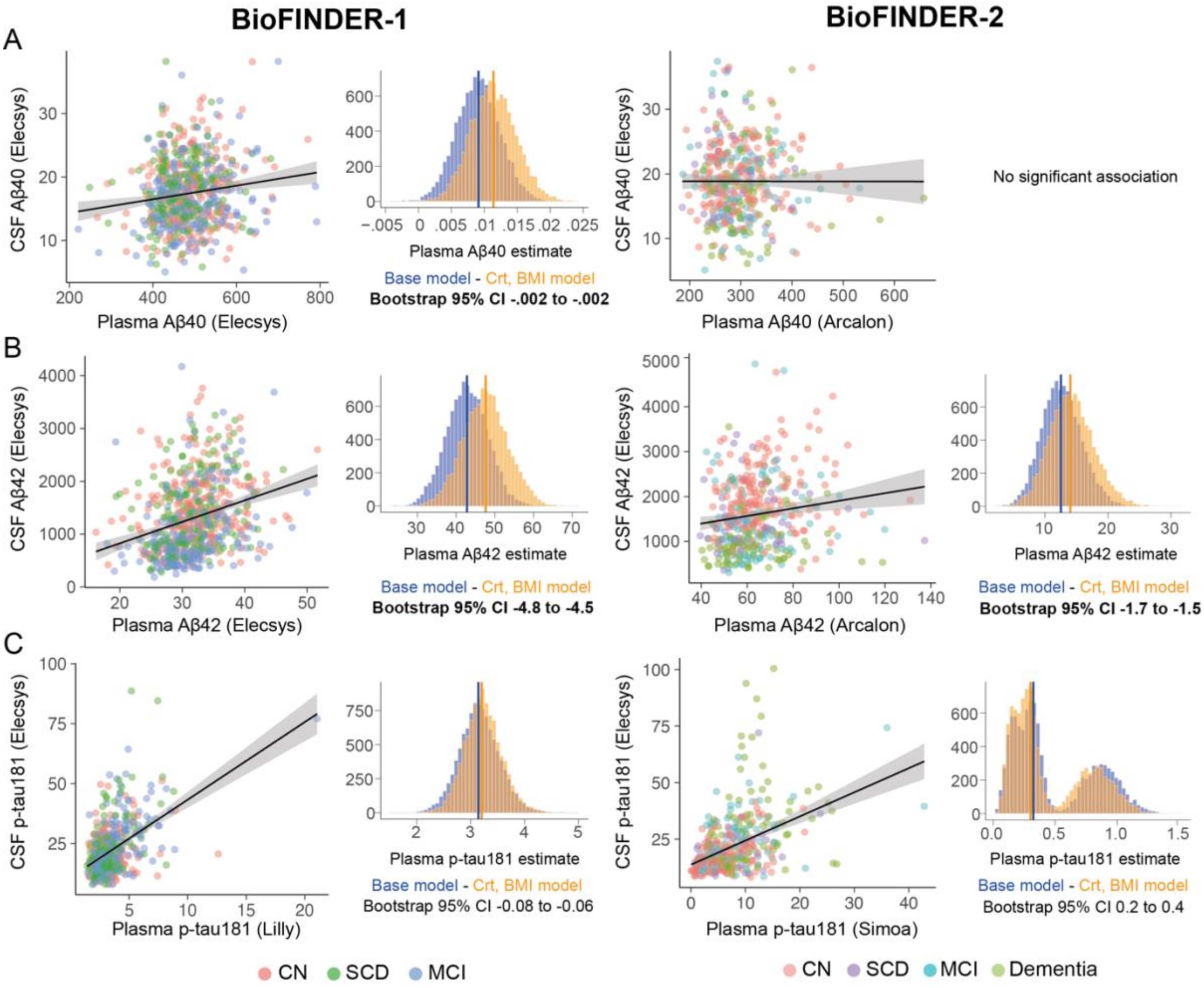


Scatter plots depict bivariate associations between plasma and CSF levels for Aβ40 (A), Aβ42 (B) and p-tau181(C) in BioFINDER-1 on the left-hand side and BioFINDER-2 on the right-hand side. To compare plasma coefficients between the base model and the one including creatinine and BMI as covariates, we generated 10 000 bootstrap samples of both models, shown in the histograms. Significant difference between models was based on the 95% confidence interval difference of the difference of plasma estimates between models. In BioFINDER-2 the association between plasma Aβ40 and CSF Aβ40 was not significant and thus comparisons between models was not tested.

Abbreviations: Aβ, beta-amyloid; BMI, body mass index; Crt: creatinine, CSF, cerebrospinal fluid, GFAP, p-tau181, phosphorylated tau 181

**Table S3. WashU-IP-MS plasma Aβ markers coefficients related to corresponding cerebrospinal fluid biomarker levels**

| | **Aβ42/40** | | **Aβ40** | | **Aβ42** | |
|---|---|---|---|---|---|---|
| | β | $R^2$ | β | $R^2$ | β | $R^2$ |
| **BioFINDER-1** | | | | | | |
| Bivariate model | **0.60** | 0.35 | **0.12** | 0.03 | **0.34** | 0.12 |
| Model including creatinine and BMI | **0.61** | 0.37 | **0.14** | 0.04 | **0.41** | 0.13 |

Standardized beta coefficients of plasma biomarkers from models assessing CSF (dependent variable) and plasma biomarkers levels adjusted for age and sex, or from models further including creatinine and BMI as covariates. Adjusted $R^2$ of models are also reported. Bolded values indicate when significant improvement in plasma estimate was seen in models including creatinine and BMI based on bootstrapping. Significance of all plasma coefficients and of all models is $p < 0.001$

Abbreviations: Aβ, beta-amyloid; BMI, body mass index

**Table S4. Plasma Aβ42 and p-tau181 estimates related to conversion to dementia from logistic regressions in BioFINDER-1**

| | **Aβ42 (Elecsys)** | | **p-tau181** | |
|---|---|---|---|---|
| | OR | AUC | OR | AUC |
| **Conversion to AD-dementia within 4 years** | | | | |
| Base model | 0.53 | 0.68 | **3.11** | **0.81** |
| Model including creatinine and BMI | 0.49 | 0.71 | **3.26** | **0.82** |
| **Conversion to all-cause dementia within 4 years** | | | | |
| Base model | 0.79 | 0.61 | 2.06 | 0.70 |
| Model including creatinine and BMI | 0.76 | 0.64 | 2.10 | 0.70 |

Odds ratio of plasma biomarkers from logistic regression to discriminate participants who remained stable vs. those who converted to AD dementia or all-cause dementia. Odds ratio represents the increased odds of converting to dementia for each increase in standard deviation biomarker value. Models included age and sex as covariates. Bolded values indicate when significant improvement in plasma estimate was seen in models including creatinine and BMI based on bootstrapping. Significance of all plasma odds ratio is $p < 0.001$, except for Aβ42 on conversion to all-cause dementia where $p=0.02$

Abbreviations: Aβ, beta-amyloid; AUC, area under the curve; BMI, body mass index; p-tau181, phosphorylated tau 181

**Figure S5. Effect of adjusting p-tau217 and NfL models for creatinine and BMI in relation to conversion to dementia at the individual level**

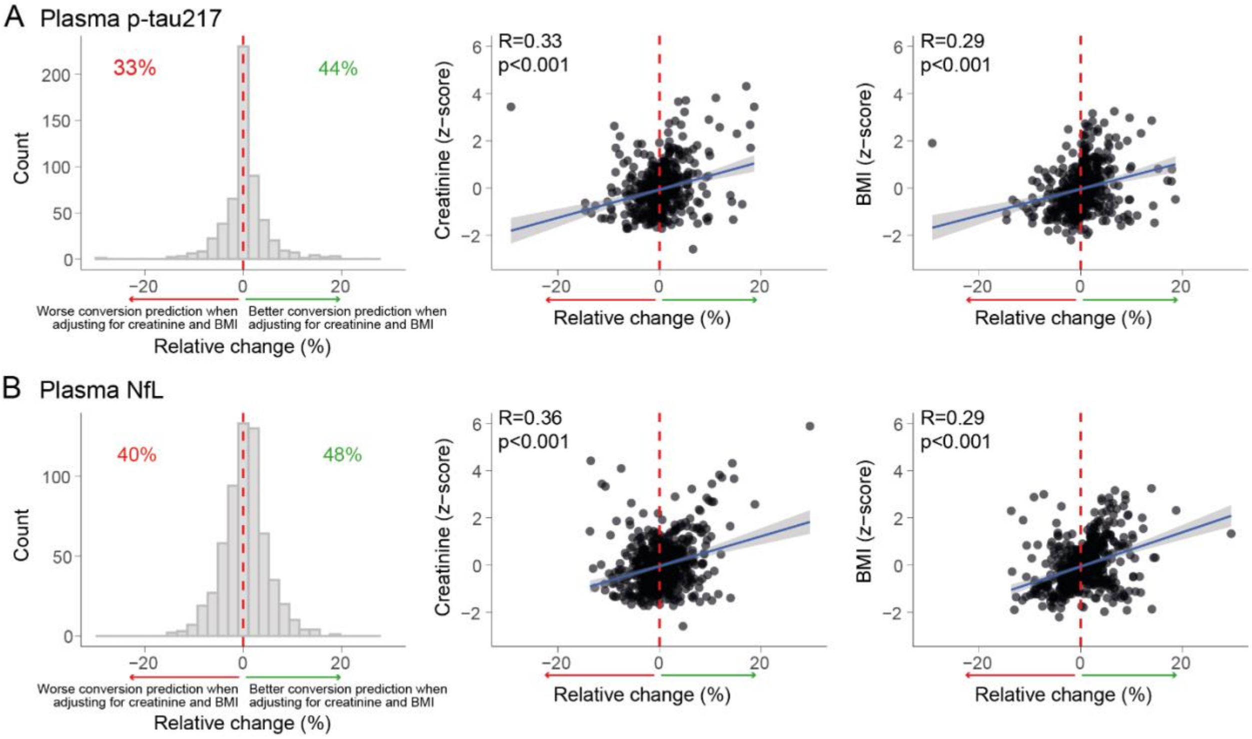


Relative change in plasma p-tau217 odds ratio related to conversion to AD dementia (A) and plasma NfL odds ratio related to conversion to all-cause dementia (B) Relative change at the individual level was calculated in a similar way as explained in Figure S3. As such, positive change (right to the 0% dotted red line) favors the model including creatinine and BMI (better prediction of conversion to dementia). Conversely, a negative change (left to the 0% dotted red line) favors the basic model (better prediction of conversion to dementia without including creatinine and BMI). Relations between relative change at the individual level and creatinine levels or BMI are shown in scatter plots. Abbreviations: BMI, body mass index; NfL, neurofilament light; p-tau217, phosphorylated tau 217
